# A phenome-wide association study identifying risk factors for pediatric post-concussion syndrome

**DOI:** 10.1101/2020.07.17.20155895

**Authors:** Aaron M. Yengo-Kahn, Natalie Hibshman, Christopher M. Bonfield, Eric S. Torstenson, Katherine A. Gifford, Daniil Belikau, Lea K. Davis, Scott L. Zuckerman, Jessica K. Dennis

## Abstract

**Objective:** To identify risk factors and generate hypotheses for pediatric post-concussion syndrome (PCS) using a phenome-wide association study (PheWAS).

**Methods:** A PheWAS (case-control) was conducted following the development and validation of a novel electronic health record-based algorithm that identified PCS cases and controls from an institutional database of >2.8 million patients. Cases were patients ages 5-18 with PCS codes or keywords identified by natural language processing of clinical notes. Controls were patients with mild traumatic brain injury (mTBI) codes only. Patients with moderate or severe brain injury were excluded. All patients used our healthcare system at least three times 180 days before their injury. Exposures included all pre-injury medical diagnoses assigned at least 180 days prior.

**Results:** The algorithm identified 274 pediatric PCS cases (156 females) and 1,096 controls that were age and sex matched to cases. Cases and controls both had a mean of >8 years of healthcare system use pre-injury. Of 202 pre-injury medical, four were associated with PCS after controlling for multiple testing: headache disorders (OR=5.3; 95%CI 2.8-10.1;*P=*3.8e-7), sleep disorders (OR=3.1; 95%CI 1.8-5.2; *P=*2.6e-5), gastritis/duodenitis (OR=3.6, 95%CI 1.8-7.0;*P=*2.1e-4), and chronic pharyngitis (OR=3.3; 95%CI 1.8-6.3;*P=*2.2e-4).

**Conclusions:** These results confirm the strong association of pre-injury headache disorders with PCS and provides evidence for the association of pre-injury sleep disorders with PCS. An association of PCS with prior chronic gastritis/duodenitis and pharyngitis was seen that suggests a role for chronic inflammation in PCS pathophysiology and risk. These factors should be considered during the management of pediatric mTBI cases.

## INTRODUCTION

The majority of traumatic brain injury (TBI) affecting children are mild traumatic brain injuries (mTBI).^1,2^ While the definition of concussion stipulates negative neuroimaging findings, the symptoms and recovery of concussions and mTBIs are indistinguishable.^3^ Although most concussions self-resolve in one to three weeks,^4^ approximately 10-30% of patients experience persistent cognitive, somatic, and emotional symptoms known as post-concussion syndrome (PCS).^3,5–7^

Despite significant efforts to understand which patients are at risk for PCS,^*8*^ PCS has no clear unifying pathophysiology.^9^ Growing evidence suggests a higher risk of PCS is associated with younger age,^10,11^ female sex,^10,12^ prior mTBI,^13,14^ and certain pre-existing conditions such as headaches,^15,16 17^ and psychiatric diagnoses.^13,17–20^ Nonetheless, models of mTBI recovery remain only modestly predictive,^17,21^ highlighting the need for further research into PCS and inclusion of previously understudied risk factors. Moreover, most previous research has been restricted to a small or moderate number of mTBI patients drawn from specialty concussion clinics ^5,13,22^ or from a single institution emergency department,^23^ with drop-out rates as high as 40%.^21^

Electronic health record (EHR)-wide investigations utilizing algorithm-based case selection offer at least two major advantages in the search for PCS risk factors. First, large numbers of patients from across the health care system can be ascertained at a fraction of the time and cost of a manually or prospectively ascertained sample,^24^ with drop-outs minimized by studying health system use patterns in the EHR, as opposed to requiring active patient engagement with the study team. Second, the entire EHR of mTBI patients can be mined using a Phenome-wide Association Study (PheWAS) to reveal pre-injury medical diagnoses that increase the risk of poor recovery.^25^

The objective of this study was to identify risk factors and generate hypotheses for pediatric PCS from across the EHR using a PheWAS, harnessing a novel EHR-based algorithm for PCS cases and controls.

## METHODS

### Setting and Study Design

A case-control study of pediatric PCS patients was undertaken with age- and sex-matched mTBI controls receiving care at Vanderbilt University Medical Center (VUMC). VUMC is a tertiary care center that provides inpatient and outpatient care in Nashville, TN. The VUMC EHR was established in 1989 and includes data on billing codes (hereafter referred to simply as codes) from the International Classification of Diseases, 9th and 10^th^ editions (ICD9 and ICD10), Current Procedural Terminology (CPT) codes, laboratory values, reports, and clinical notes. The de-identified mirror of the EHR used for research includes patient records on more than 2.8 million individuals. Data analyzed in this study spanned May, 1989 to March, 2019.

### Standard Protocol Approvals, Registrations and Patient Consents

The VUMC Institutional Review Board oversees the VUMC EHR research database and approved this study (IRB 151116). The research was performed without patient involvement and relied upon deidentified data for which patient consent was waived.

### Case Selection Algorithm Development and Validation

The present study of pediatric PCS patients is part of a broader research program on using EHRs to study PCS, in which we designed and evaluated four different strategies for identifying PCS cases from the EHR (Figure 1). First, we excluded patients with evidence of moderate or severe TBI, which we defined as a codes mapping to the categories of skull fracture, contusion, or hemorrhage, or a neurosurgical procedure code (ICD9: 01, 02; ICD10: 00, ON, R40.242, R40.243; CPT: 61000-62258).^26^ Next, we specified four case definitions that selected patients based on the presence of PCS codes (ICD9: 310.2; ICD10: F07.81) and/or keywords in outpatient clinical notes (“postconcuss” or “post-concuss” or “post concuss”). Case Definition 1 required patients to have two PCS codes separated by at least 14 days, an ascertainment strategy that was successful in previous analyses.^24^

**Figure 1.**
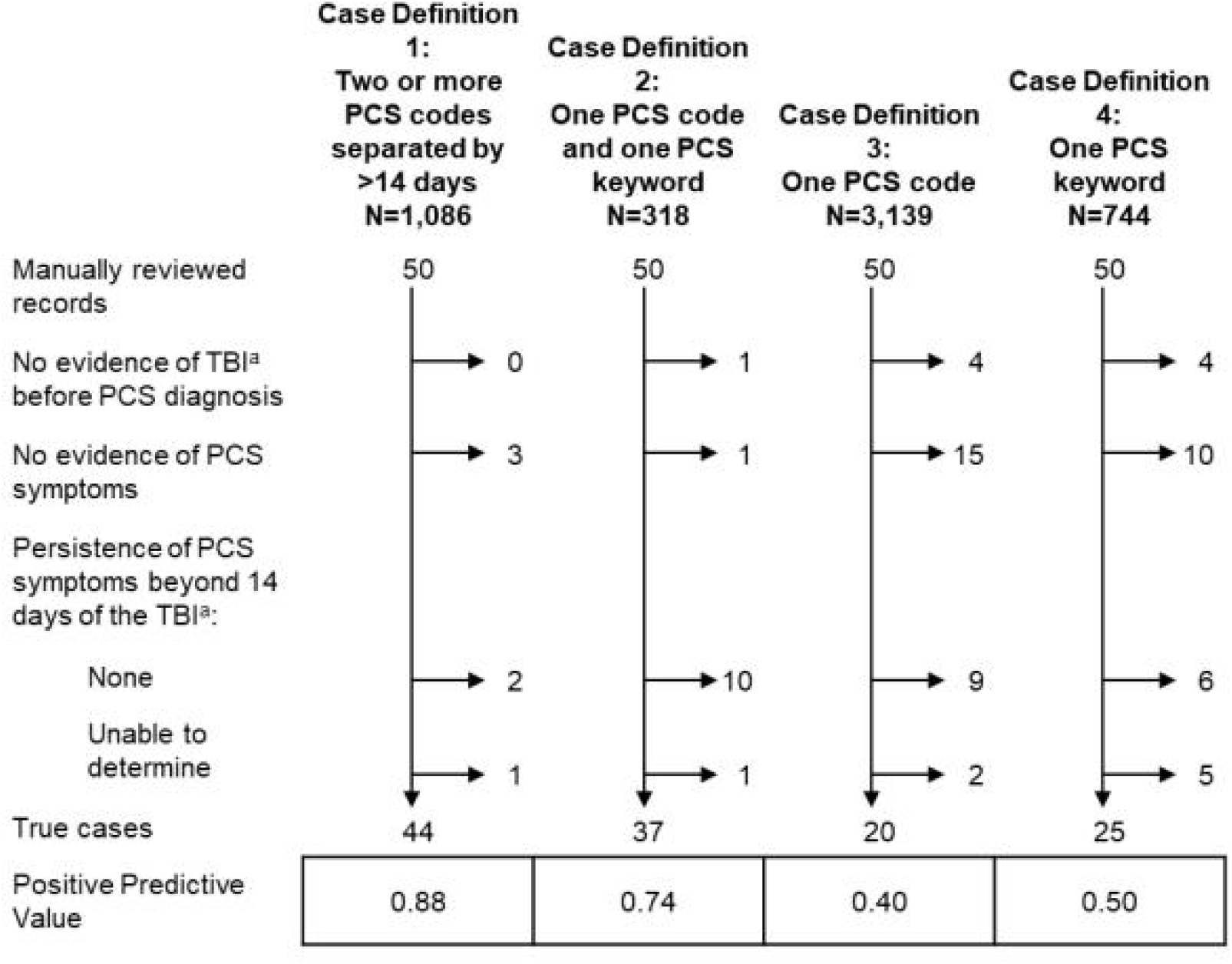
Evaluation of strategies to ascertain post-concussion syndrome (PCS) cases from electronic health records. TBI denotes traumatic brain injury and PCS, post-concussion syndrome.

Case Definition 2 required patients to have one PCS code, as well as one PCS keyword, which was identified by applying natural language processing to the outpatient clinical notes in three stages. First, outpatient clinical notes containing PCS keywords using the crude regular expression “post[-]?concuss”, which covers the keywords “postconcuss”, “post-concuss”, and “post concuss,” were queried and aggregated. Second, the records were trimmed down to sentences containing PCS keywords using the natural language processing (NLP) library spaCY.^27^ Third, these sentences were evaluated for negation terms (e.g., “no evidence of postconcussion syndrome”; “postconcussion syndrome not suspected”). For this, the library pyConTextNLP^28^ was utilized and assigned weights to each sentence corresponding to the evidence in support of PCS. Weights were: (0) sentences contained conflicting or no definitive evidence such as entries in a problem list; (1) sentences were suggestive of positive evidence; (2) sentences had definitively positive evidence; (−1) sentences were suggestive of negative evidence; (−2) sentences had definitively negative evidence. Weights were summed across PCS containing sentences in the clinical note and a final score was assigned to the note itself. Clinical notes with scores of 0 or more were considered to have evidence of PCS.

Case Definition 3 required patients to have only one PCS code, while Case Definition 4 required patients to have only one PCS keyword. Across all case groups, we only included cases whose last PCS mention (code or keyword) occurred when they were five years of age or older, since symptoms reported in children younger than five years old are difficult to confirm.^29^

We evaluated each case definition by manually reviewing the EHRs of 50 randomly selected patients from each group, operationally defining PCS as persistent symptoms for longer than two weeks.^4,30^ We abstracted data on the mTBI preceding the PCS diagnosis, including information on care sought, brain imaging, and injury cause, as well as persistence of >1 PCS symptom, as defined as any of the 22 symptoms listed on the PCS symptom scale contained within the 5th edition of the sport concussion assessment tool (SCAT5).^3^ Data were abstracted into a REDCap database by one reviewer (NH), and difficult-to-classify data elements were resolved by consensus (NH, JKD, and AYK). Patients were classified as true positives if they had both evidence of a mTBI preceding the PCS diagnosis, and if PCS symptoms persisted beyond 14 days of the mTBI.^4^ We calculated the positive predictive value (PPV) of each case definition as the number of true cases divided by the number of cases meeting the case definition.

Manual Review identified 44 and 37 true positives, yielding PPVs of 88% and 74% respectively for case definitions 1 and 2 (Figure 1). The PPVs of case definitions 1 and 2 were 100% and 64% respectively in the subset of manually reviewed pediatric patients, and descriptive characteristics of the true positive set are presented in Table 1.

**Table 1:** Characteristics of the true positives identified by manual review of 100 records meeting case definitions 1 and 2.

|  | True Positives (N=81) | Pediatric True Positives (N=13) |
| --- | --- | --- |
| Care sought for index TBI |  |  |
| Yes – VUMC | 39 (48.1) | 8 (61.5) |
| Yes – Not VUMC | 21 (25.9) | 3 (23.1) |
| No | 1 (1.2) | 0 (0) |
| Unknown | 20 (24.7) | 2 (15.4) |
| Hospital length of stay for index TBI |  |  |
| None – ED visit | 41 (50.6) | 9 (69.2) |
| None – outpatient visit | 3 (3.7) | 1 (7.7) |
| Hospitalization <3 days | 2 (2.5) | 0 (0) |
| Hospitalization ≥ 3 days | 4 (4.9) | 1 (7.7) |
| Unknown | 31 (38.3) | 2 (15.4) |
| Imaging for the index TBI | 55 (67.9) | 10 (76.9) |
| Positive imaging result | 8 (9.9) | 1 (7.7) |
| Cause of index TBI |  |  |
| Motor vehicle collision | 35 (43.2) | 4 (30.8) |
| Fall – not sports-related | 29 (35.8) | 2 (15.4) |
| Assault | 5 (6.2) | 1 (7.7) |
| Sports-related | 6 (7.4) | 5 (38.5) |
| Other / unknown | 6 (7.4) | 1 (7.7) |

### Selection of PheWAS Participants

Analyses proceeded with cases identified with case definitions 1 and 2 given superior PPV (Figure 1). Each PCS case was matched to four controls on sex and age (within one-year intervals). Controls had no PCS codes or keywords in their EHR, but evidence of an mTBI, i.e. codes that mapped to the categories of concussion / loss of consciousness, or other and unspecified head injury.^26^ Patients with evidence of a moderate or severe TBI were excluded from both groups.

In both cases and controls, an index event was defined to delineate pre-injury risk factors for PCS. In cases, the index event was the first mTBI code, or the first PCS code if the patient had no mTBI code or if the first PCS code preceded the first mTBI code. In controls, the index event was the first mTBI code. To ensure adequate data for PheWAS analyses, patients had to have visited VUMC at least three times at least 180 days before the index event. Additionally, controls had to have visited VUMC at least once 30 days after the index event, to ensure that they continued to use VUMC post-injury and had not sought PCS care elsewhere. Finally, we excluded patients whose index event occurred over age 18 or later. In the primary analysis, ICD codes were restricted to those assigned at least 180 days before the index event, identifying diagnoses long preceding the injury.

### Exposure Assignment

Phecodes are used in PheWAS to collapse multiple ICD9 and ICD10 codes into a smaller number of higher-order disease categories. The Phecode Map v1.2(beta), as implemented in the PheWAS R package v0.99.5-2,^31^ was used to map ICD9 and ICD10 codes in this >180 day window to phecodes. Patients were assigned to the affected group for a given phecode if they had at least one ICD code that mapped to a given phecode, otherwise they were assigned to the unaffected group. Although most PheWAS analyses in adults require at least two ICD codes before assigning a patient to an affected phecode group,^25^ the threshold was lowered to one ICD code to account for the lighter health system usage among children.

### Statistical Analyses

Descriptive characteristics of the matched cases and controls were compared using a conditional logistic regression model to account for matching. Statistically significant differences existed in the pre-injury time spent in the EHR and number of pre-injury ICD codes, thus these variables were included as covariates in the PheWAS analysis. PCS case/control status was modeled as the outcome variable and each phecode was considered a predictor variable in separate conditional logistic regression models (clogit function in the R survival package). To ensure sufficient cell counts for interpretable odds ratios, only phecodes with at least 25 affected patients were included. In the primary analysis, 201 phecodes met this threshold and the Bonferroni correction for phenome-wide statistical significance was *P*<2.48e-4 (0.05/201). Sex-stratified analysis on the four phecodes that passed our threshold for statistical significance was performed and the effect size of these phecodes between sexes was compared.

Three additional sensitivity analyses were conducted to assess the robustness of the results: (1) restricting PheWAS to the cases and matched controls identified by case definition 1, which had the strongest positive predictive value (PPV); (2) restricting PheWAS to the cases and matched controls with a documented mTBI in their EHR; and (3) broadening the pre-injury window to include ICD codes diagnosed at least 30 days before the index event (as opposed to 180 days in the primary analysis). The pre-injury window was not broadened to include codes assigned immediately before or at the same time as the index event because the focus was on pre-injury risk factors, which have been incompletely documented in previous research. All statistical analyses were performed in R 3.6.0 (R Foundation, Vienna, Austria). There were no missing data in the primary analysis and no data were imputed.

### Data Availability Statement

Full results are available in a public, open access repository (https://dennislab.ca/resources-and-software/). Data may be used with proper citation and crediting.

## RESULTS

### Participant Characteristics

PheWAS selection criteria identified 274 pediatric cases and 9,069 potential pediatric controls (Figure 2). Cases were older than eligible controls (median age 15.0 vs. 12.1) and were more likely to be female (56.9% vs. 39.1%). Therefore, each case was matched on age and sex to four eligible controls, for a final sample size of 274 cases (156 females and 118 males) and 1096 controls (Table 2). Of these 274 cases, 196 (71.5%) were identified by case definition 1. Over three-quarters of cases had a documented mTBI in their EHR.

**Table 2:** Descriptive characteristics of the matched sample used in the PheWAS.

| Participant characteristics<sup>a</sup> | Controls (N=1096) | Cases (N=274) | *P* value<sup>b</sup> |
| --- | --- | --- | --- |
| Age at index event in years | 14.4 (3.1) | 14.4 (3.1) | NA |
| Female, N (%) | 624 (56.9) | 156 (56.9) | NA |
| Number of days in EHR before index event | 2929 (1671.2) | 3302 (1637.3) | 8.6e-4 |
| Number of ICD codes at least 180 days before index event | 34.0 (68.6) | 44.5 (101.6) | 0.058 |
| Number of visits at least 180 days before index event | 19.2 (33.7) | 20.6 (31.3) | 0.550 |
| Number of ICD codes at least 30 days before index event | 37.0 (73.8) | 50.2 (114.1) | 0.038 |
| Number of visits at least 30 days before index event | 20.8 (35.0) | 22.5 (33.8) | 0.480 |
| Documented TBI in EHR, N (%) | 1096 (100) | 208 (75.9) | NA<sup>c</sup> |
| Motor vehicle collision cause code<sup>d</sup> before index event, N (%) | 107 (9.8) | 6 (2.2) | 1.2e-4 |

<sup>a</sup>Data are presented as mean (sd) unless noted otherwise, <sup>b</sup>*P* values were calculated from a conditional logistic regression model, <sup>c</sup>Model failed to converge due to a complete separation of the data, <sup>d</sup>ICD9 and ICD10 external cause of injury codes in patients' EHRs were mapped to the motor vehicle cause of injury category as previously published.<sup>26</sup>

**Figure 2.**
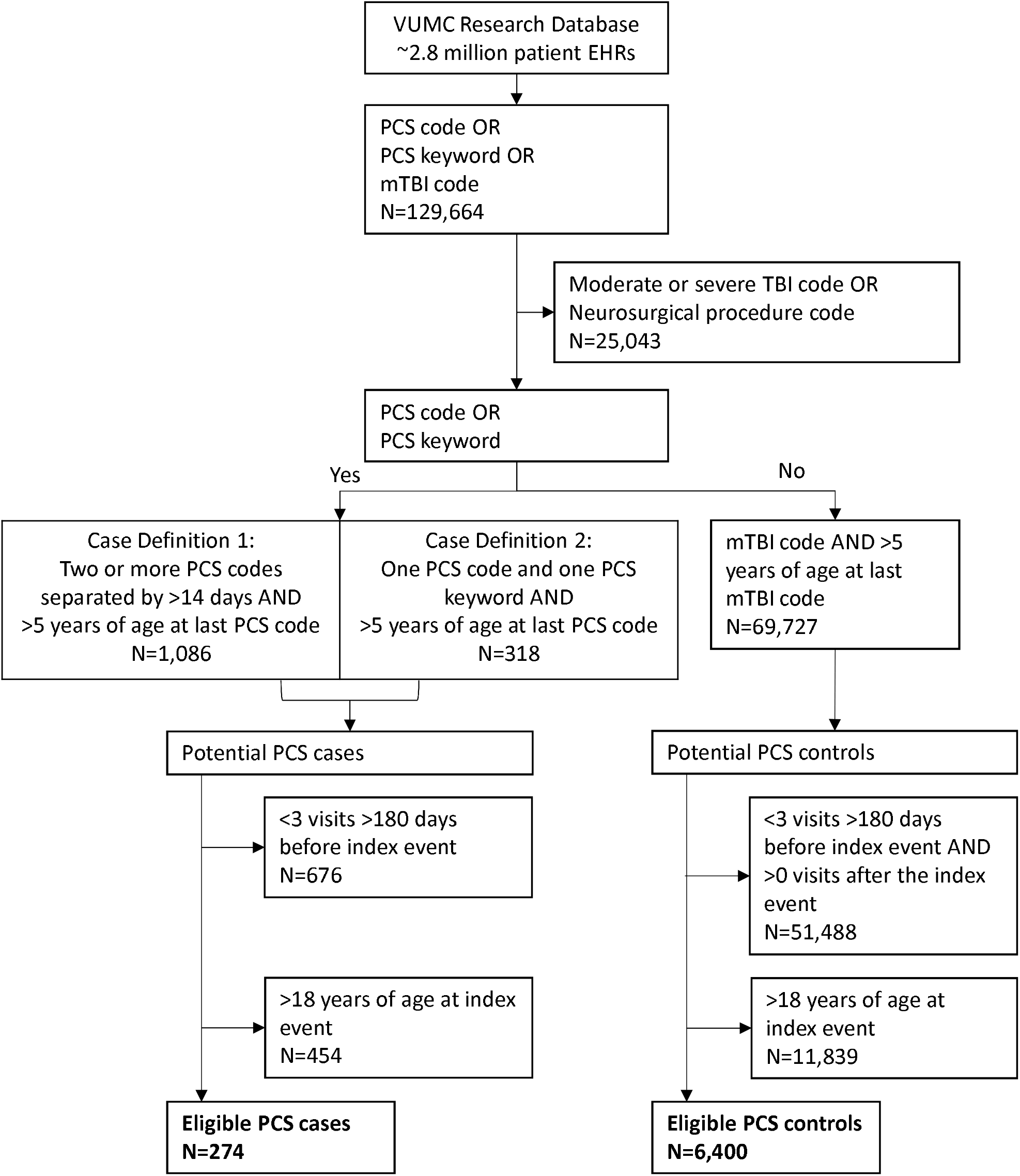
Patient Selection Flow Diagram. In controls, the index event was the first mTBI code. In cases, the index event was the first mTBI code, or the first PCS code if there was no mTBI code assigned or if the first mTBI code occurred after the first PCS code. EHR denotes Electronic health records; PCS, post-concussion syndrome, PPV, positive predictive value; (m)TBI, (mild) traumatic brain injury; and VUMC, Vanderbilt University Medical Center.

Within the matched sample (Table 2), cases and controls had a comparable number of visits at least 180 days before the index event (20.6 vs. 19.2, *P=*0.550). Cases, however, had a higher number of codes assigned at least 180 days before the index event (44.5 vs 34.0, *P=*0.058) and a higher number of days in the EHR pre-index event (3302 days in cases vs. 2929 days in controls,*P=*8.6e-4). Though both groups were in the EHR long before the index injury (>8 years), to safeguard against group differences, the PheWAS analyses included covariates for the number of codes and days pre-index event.

### PheWAS

In the primary analysis of codes assigned at least 180 days before the index event, cases were more likely than controls to have reported headache syndromes (OR=5.3; 95% CI 2.8-10.1; *P=*3.8e-7), sleep disorders (OR=3.1; 95% CI 1.8-5.2; *P=*2.6e-5), gastritis and duodenitis (OR=3.6, 95% CI 1.8-7.0; *P=*2.1e-4), and chronic pharyngitis and nasopharyngitis (OR=3.3; 95% CI 1.8-6.3; *P=*2.2e-4)(Figure 3;Table 3). Cases were also more likely than controls to have a pre-injury diagnosis of anxiety disorder (OR=2.4; 95% CI 1.4-4.2; *P=*0.001). This phecode fell below the threshold for phenome-wide significance but was the most strongly associated psychiatric phecode. Phecode associations were similar in males and females (Table 3). Full sex-stratified PheWAS results are shown graphically in Supplemental Figures 1 and 2.

**Table 3:** Phecodes associated with pediatric PCS at the phenome-wide threshold for statistical significance.

| Phecode | Overall |  |  |  | Females |  |  |  | Males |  |  |  | Females<br>vs. Males |
| --- | --- | --- | --- | --- | --- | --- | --- | --- | --- | --- | --- | --- | --- |
|  | N<br>True | N<br>False | OR (95% CI) | P value | N<br>True | N<br>False | OR (95% CI) | P value | N<br>True | N<br>False | OR (95% CI) | P value | P value |
| Other headache syndromes | 41 | 1329 | 5.3 (2.8-10.1) | 3.85e-7 | 32 | 748 | 4.3 (2.1-8.6) | 7.53e-5 | 9 | 581 | 12.4 (2.5-60.6) | 0.002 | 0.84 |
| Sleep disorders | 62 | 1308 | 3.1 (1.8-5.2) | 2.60e-5 | 32 | 748 | 4.1 (1.9-8.7) | 2.37e-4 | 30 | 560 | 2.3 (1.1-5.0) | 0.028 | 0.26 |
| Gastritis and duodenitis | 44 | 1326 | 3.6 (1.8-7.0) | 2.08e-4 | 26 | 754 | 3.8 (1.6-9.0) | 0.003 | 18 | 572 | 3.4 (1.2-10.0) | 0.024 | 0.46 |
| Chronic pharyngitis and nasopharyngitis | 44 | 1326 | 3.3 (1.8-6.3) | 2.21e-4 | 36 | 744 | 4.3 (2.1-8.8) | 9.54e-5 | 8 | 582 | 1.3 (0.3-6.4) | 0.756 | 0.14 |
| Anxiety disorders<sup>b</sup> | 71 | 1299 | 2.4 (1.4-4.2) | 0.001 | 51 | 729 | 1.7 (0.9 - 3.4) | 0.131 | 20 | 570 | 4.7 (1.9 - 11.8) | 0.001 | 0.87 |

<sup>a</sup>The phecode “Other headache syndromes” includes all headaches other than tension and migraine headaches.
<sup>b</sup>The association between “Anxiety disorders” and PCS fell just below the threshold for phenome-wide significance. “Anxiety disorders”, however, were the fifth-ranked phecode in the PheWAS and results are included in this table for comparison with previous literature.

**Figure 3.**
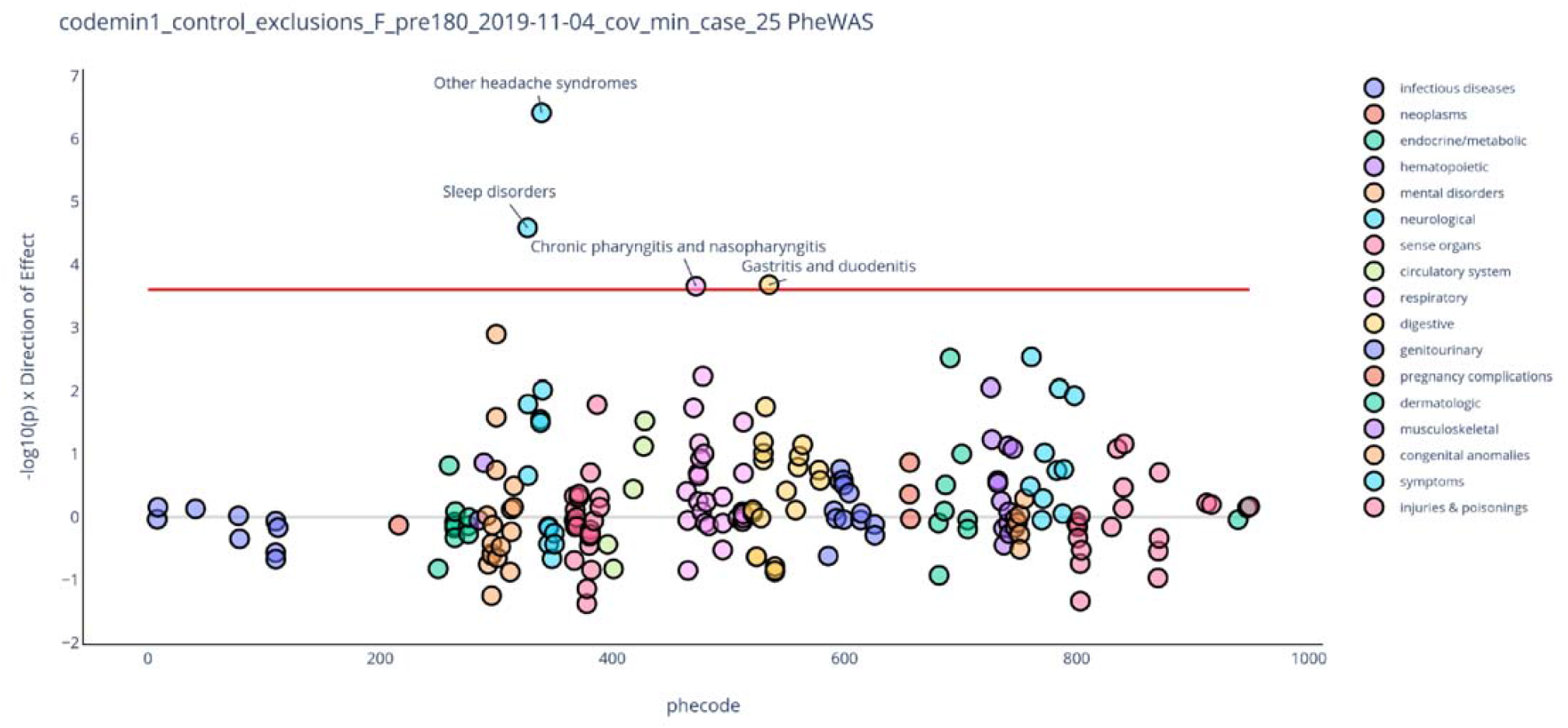
PheWAS of 201 Phecodes Assigned ≥180 Days Pre-Index Event. To ensure sufficient cell counts for interpretable odds ratios, only phecodes with at least 25 affected patients were included. The red line denotes the Bonferroni threshold for phenome-wide significance (0.05/201=2.48×10-4). Phecodes plotted above the x-axis are associated with an increased odds of PCS while those plotted below the x-axis are associated with a reduced odds of PCS. Interactive plots and full results are available at [https://dennislab.ca/resources-and-software/].

Restricting analysis to the 196 cases ascertained using case definition 1, results were similar to the primary analysis, with the exception of an attenuated effect estimate for other headache syndromes (OR 1.9; 95% CI 0.68-5.17; *P=*0.221)(Table 4). Restricting analysis to the 208 cases with a documented mTBI in their EHR did not appreciably change any of the top phecode associations (Table 4). Using phecodes diagnosed at least 30 days before the index event, results were largely unchanged compared to those in the primary analysis, except that the migraine phecode now crossed the threshold for phenome-wide significance (OR 2.5; 95% CI 1.6-4.1; *P=*1.7e-4)(Supplemental Figure 3).

**Table 4:** Sensitivity analysis of top phecodes associated with pediatric PCS.

| Phecode | Including only cases meeting case definition 1 |  |  |  | Including only cases with a traumatic brain injury code |  |  |  |
| --- | --- | --- | --- | --- | --- | --- | --- | --- |
|  | N True | N False | P value | OR (95% CI) | N True | N False | P value | OR (95% CI) |
| Other headache syndromes | 18 | 962 | 0.221 | 1.9 (0.7 - 5.2) | 26 | 864 | 3.86E-04 | 4.3 (1.9 - 9.5) |
| Sleep disorders | 45 | 935 | 3.26E-04 | 3.1 (1.7 - 5.9) | 38 | 852 | 2.73E-02 | 2.2 (1.1 - 4.3) |
| Gastritis and duodenitis | 27 | 953 | 0.004 | 3.4 (1.5 - 7.7) | 26 | 864 | 5.42E-04 | 4.7 (2.0 - 11.5) |
| Chronic pharyngitis and nasopharyngitis | 30 | 950 | 0.046 | 2.2 (1.0 - 4.9) | 23 | 867 | 0.010 | 3.4 (1.3 - 8.5) |
| Anxiety disorder | 47 | 933 | 0.011 | 2.4 (1.2 - 4.5) | 52 | 838 | 0.002 | 2.8 (1.5 - 5.3) |

## DISCUSSION

Our EHR-based PCS algorithm identified 274 pediatric PCS cases and 1096 matched pediatric mTBI controls from a single health care system in the United States. We then leveraged the entire EHR of cases and controls (8-9 years pre-injury, on average) to test over 200 pre-injury diagnoses for association with PCS risk using a PheWAS. Results from the PheWAS recapitulated established PCS risk factors (headache and migraine history) and identified previously underreported associations with pre-injury sleep disorders and inflammatory-related diagnoses.

Pediatric mTBI is a major public health concern, especially for the subset of children who develop PCS. Several governing bodies, including the Centers for Disease Control and Prevention, have called for more research to improve our understanding of mTBI and its sequelae, specifically addressing the small sample sizes and biases in existing research.^32^ Our EHR-based study directly responds to these research gaps. First, our PCS algorithm identified among the largest samples of PCS cases and controls reported to date,^8^ and is portable to other EHR-based health care systems, facilitating future large-scale studies of pediatric mTBI recovery. Second, our study included a broad mTBI population (all modes of injury recruited from across the healthcare system) for maximum generalizability. Third, we required patients to have used our health care system a minimum number of times before and after the index event to help ensure that cases and controls used VUMC for all their medical care. This requirement meant that we captured mTBI patients with favorable outcomes and included them as controls, whereas prospective studies of mTBI patients often lose these patients to follow-up.^21^ Finally, all objectively measured medical diagnoses before the index event were included as predictor variables, avoiding our own biases in selecting candidate risk factors, as well as participants’ recall biases.

Pre-injury headache is a consistent risk factor for PCS.^8^ In a prospective multi-center analysis of 2,006 pediatric concussion patients, a self-reported physician-diagnosed history of migraines nearly doubled the risk of PCS at 28 days.^17^ Another large retrospective cohort study of nearly 2,000 pediatric sport-related concussion patients demonstrated that self-reported pre-morbid headaches statistically increased the risk of prolonged recovery by 15%.^16^ In comparison, we found that other headache syndromes and migraines diagnosed by a physician and recorded in the EHR at least 180 days pre-injury increased the risk of PCS by two- to five-fold. Our effect estimates are higher than those previously reported, which could be due to differences in study design. We included a broad pediatric mTBI population as opposed to only sport-concussion patients included in previous studies, and pre-injury medical diagnoses were objectively ascertained instead of by self-report.

Numerous investigations over the past decade have demonstrated a relationship between post-mTBI sleep disturbances and prolonged recovery,^33^ increased symptom reporting,^34^ and worsened neurocognitive test scores.^34,35^ Another study recently demonstrated that 78% of adult patients diagnosed with PCS who underwent a sleep study after the index head injury were diagnosed with sleep apnea.^36^ However, only one study ^37^ has evaluated the effect of pre-mTBI sleep difficulties in an older group of 348 adolescent and adult athletes and found that the 34 athletes with self-reported pre-injury sleeping difficulties demonstrated worse reaction times and higher symptom scores at 2, 5-7 and 10-14 days post-injury. Unfortunately, the study did not determine whether those with sleep difficulties went on to develop PCS. We systematically assessed physician-diagnosed pre-morbid sleep disorders and show these are a risk factor for poor recovery and specifically, PCS.

The link between pre-injury sleep disorders and mTBI recovery is likely multifactorial, including decreased cognitive reserve in sleep-deprived individuals,^38^ especially those with obstructive sleep apnea.^39^ The link, however, may also be attributable to poor psychomotor performance, reaction time, and postural control in sleep-deprived individuals, leading to increased mTBI incidence and severity.^40,41^

The association between chronic inflammation and PCS is presently underreported. We identified an over three-fold increased risk of PCS associated with both previously diagnosed gastritis/duodenitis and chronic pharyngitis/nasopharyngitis. This finding may reflect frequent health system utilization among cases, but cases and controls had a comparable number of visits before the index event. Alternatively, this finding could represent underlying chronic inflammatory processes conferring risk for prolonged or poor recovery after mTBI.

Gastritis, duodenitis, pharyngitis, and nasopharyngitis are diagnoses that involve activation of peripheral immune cells such as monocytes and neutrophils.^42^ These same immune cells also cross the blood brain barrier after an TBI,^43^,^44^ and animal models have demonstrated improved functional recovery and reduced edema, microglial activation and TBI lesion size after TBI by blocking the migration of monocytes into the central nervous system,^45^ or by depleting systemic neutrophils.^46^ We therefore hypothesize that a baseline state of chronic inflammation in patients with gastritis, duodenitis, pharyngitis, and nasopharyngitis, could produce a more profound inflammatory response to mTBI and consequently, a prolonged recovery. These findings warrant investigations of inflammatory markers and cytokine profiles in mTBI recovery.

A major strength of our study is the large number of pediatric PCS cases and controls from across the health care system and with dense EHR data, which allowed testing of over 200 pre-injury medical diagnoses for association with PCS. A limitation is the imperfect PPVs of our case selection strategies. Our algorithm’s PPVs, however, are in line with the PPVs of other EHR-based algorithms,^47,48^ and most top phecode associations were similar in the PheWAS restricted to the cases identified by Case Definition 1, which had the higher PPV. The association with other headache syndromes was attenuated, although the number of patients with this phecode was greatly reduced (18 in the sensitivity analysis vs. 41 in the primary analysis). The attenuated effect estimate could also be because patients reporting chronic headache are eventually misdiagnosed as having PCS. A second potential study limitation is our definition of PCS based on symptom presence at 14 days post-injury. Some have argued that typical recovery takes longer than 14 days in children,^3^ but the definition of PCS is inconsistent across clinicians^9,30^ and diagnostic manuals,^49,50^ and a large study of 1,953 pediatric sports concussion patients reported a median time to symptom resolution of 18 days (range 1-353).^16^ A third potential limitation is heterogeneity in mTBI severity. In manual review, we found that nearly three-quarters of PCS cases sought care for the mTBI, whereas by design, all controls sought care for their injury. Nonetheless, injury severity characteristics (e.g., loss of consciousness and post-traumatic amnesia) are inconsistent predictors of PCS risk,^8^ and sensitivity analyses of the PCS cases with a documented mTBI in their EHR yielded results similar to those in the primary analysis. Fourth, our PheWAS relied on pre-injury billing code diagnoses, which can be imprecise. For example, other headache syndromes and migraine are captured by mutually exclusive billing codes and phecodes. Clinically, however, these diagnoses may overlap, and there could be some misattribution of other headache syndromes to migraines, and vice-versa. Finally, the patients utilized for the study were cared for within a single health system in the United States, which may limit the generalizability of the findings.

## CONCLUSION

An EHR-based PCS algorithm can be used to identify a broad group of pediatric mTBI patients with and without PCS, and to study pre-injury medical diagnoses associated with PCS risk in a PheWAS. The results confirm the association of pre-injury headache disorders with PCS and provide evidence for the association of pre-injury sleep disorders with PCS. An association of PCS with prior chronic gastritis/duodenitis and pharyngitis was seen, suggesting a role for chronic inflammation in PCS pathophysiology and risk. Evaluation of these factors should be considered during the clinical management of pediatric mTBI cases.

## Data Availability

https://dennislab.ca/resources-and-software/

## Appendix 1: Authors

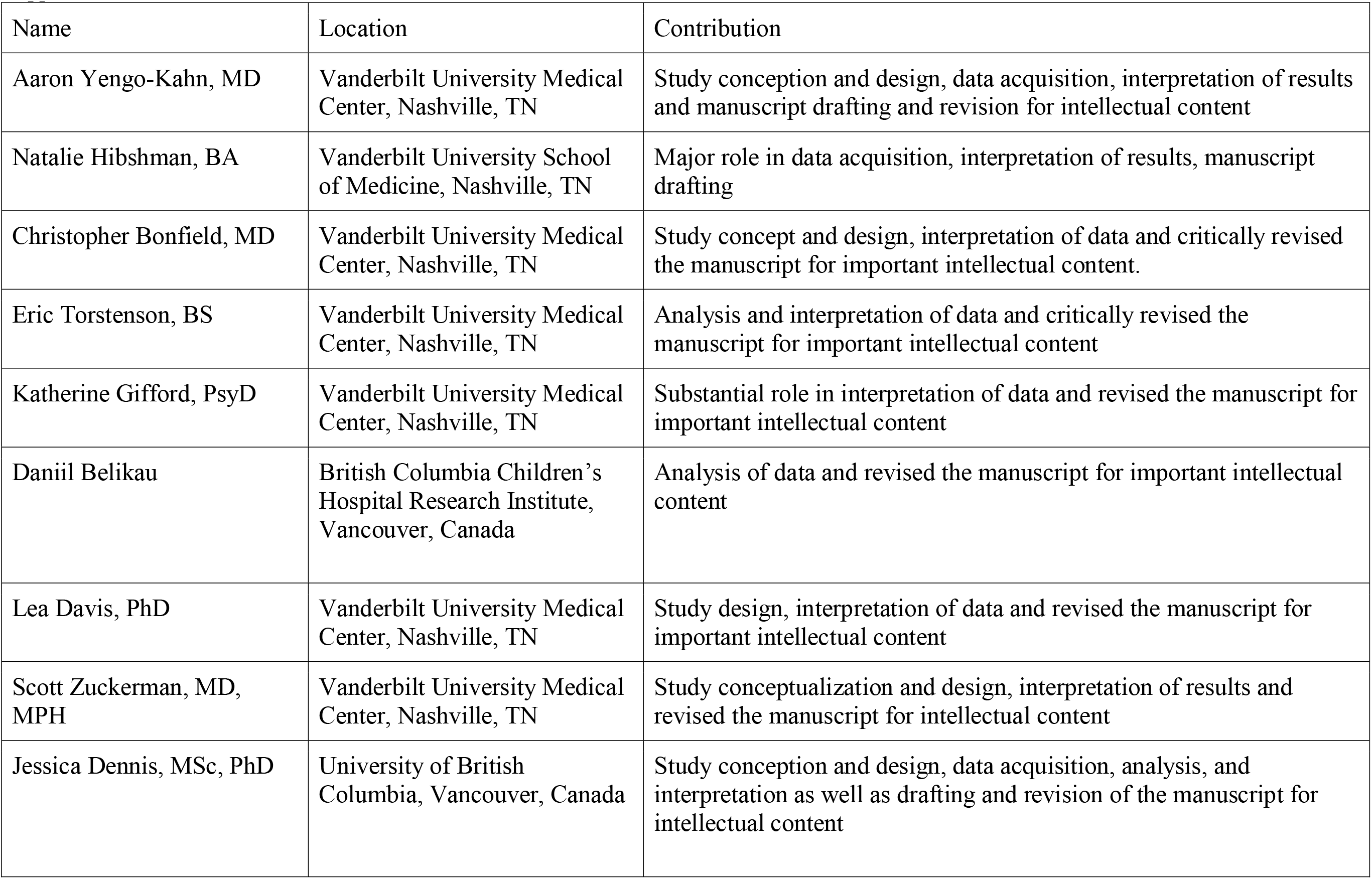

